# Using Convolutional Neural Networks for the Classification of Suboptimal Chest Radiographs

**DOI:** 10.1101/2024.11.10.24317051

**Authors:** Emily Liu, D Carrion, MK Badawy

## Abstract

**Background:** Chest X-rays (CXR) rank among the most conducted X-ray examinations. They often require repeat imaging due to inadequate quality, leading to increased radiation exposure and delays in patient care and diagnosis. This research assesses the efficacy of DenseNet121 and YOLOv8 neural networks in detecting suboptimal CXRs, which may minimise delays and enhance patient outcomes.

**Method:** The study included 3,587 patients with a median age of 67 (0-102). It utilised an initial dataset comprising 10,000 CXRs randomly divided into a training subset (4,000 optimal and 4,000 suboptimal) and a validation subset (400 optimal and 400 suboptimal). The test subset (25 optimal and 25 suboptimal) was curated from the remaining images to provide adequate variation. DenseNet121 and YOLOv8 were chosen due to their capabilities in image classification. Their performance was assessed via the area under the receiver operating curve (AUROC) and compared to radiologist classification, utilising the chi-squared test.

**Results:** DenseNet121 attained an AUROC of 0.97, while YOLOv8 recorded a score of 0.95, indicating a strong capability in differentiating between optimal and suboptimal CXRs. The alignment between radiologists and models exhibited variability partly due to the lack of clinical indications. However, the performance was not statistically significant.

**Conclusion:** Both AI models effectively classified chest X-ray quality, demonstrating the potential for providing radiographers with feedback to improve image quality. Notably, this was the first study to include both PA and lateral CXRs, as well as pediatric cases, and the first to evaluate YOLOv8 for this application.

## Introduction

Artificial intelligence (AI), a branch of computer science, focuses on creating systems capable of performing tasks traditionally executed by humans. In medical imaging, integrating visual intelligence into AI allows models to classify images based on relevant characteristics^1^. Deep learning algorithms enhance diagnostic accuracy in medical imaging by identifying abnormalities, segmenting images, and automating bone measurements^2,3^. Additionally, certain AI models can detect inaccuracies in substandard radiographs^3^.

Reject analysis plays a pivotal role in quality assurance within radiology by identifying suboptimal images and reducing the necessity for repeat examinations^4^. Chest X-rays (CXRs) are among the most performed imaging studies in clinical practice and tend to have higher rejection rates than other modalities^5^. The primary reasons for rejecting CXRs include issues with collimation and positioning, such as the scapula obscuring the lung fields, which may result from the patient’s critical condition or insufficient attention from radiographers^5,6^. Furthermore, discrepancies in quality perception between radiographers and radiologists - where radiographers base their judgments on technical criteria, potentially rejecting images that radiologists might find acceptable - contribute to image rejection rates ^5,7^. Consequently, reject analysis is essential for identifying the causes and frequency of rejected images and developing strategies to mitigate these issues^4^.

A comprehensive literature review highlights a gap in models capable of effectively assessing technical errors in suboptimal radiographs, with few solutions addressing this challenge^8,9^. This study employs AI-driven neural networks, specifically DenseNet121 and YOLOv8, to analyse CXRs, encompassing high-quality and substandard images. DenseNet121 is widely used in medical imaging due to its dense connections that enhance feature learning, making it highly suitable for complex image classification tasks^10^. YOLOv8, a state-of-the-art model, excels in feature extraction and integration, enabling rapid and precise classification^11^. The selection of these models is justified by their exceptional performance in image classification tasks^8,9,11^.

Integrating AI into radiology Quality Control (QC) has meaningful clinical advantages, such as ensuring consistent reporting, easing observer workload, and broadening quality checks across all radiographs^8^. Efficiently detecting rejects using AI enables auditing large volumes of images and providing feedback and education, thereby potentially reducing reject rates. This, in turn, can decrease radiation exposure, streamline workflow, expedite patient care and diagnosis, and enhance the overall patient experience^6^. Moreover, improving the quality of CXRs is crucial for accurately assessing critical findings such as pneumothorax, pneumonia, and lesions, thereby directly impacting patient outcomes^12^.

This study assesses whether the DenseNet121 and YOLOv8 convolutional neural networks can effectively classify CXRs as optimal or suboptimal. Notably, it is the first study to include both PA and lateral CXRs, as well as pediatric cases, and the first to evaluate YOLOv8 in this context.

## Methodology

This study utilised a quantitative, retrospective, and experimental approach to evaluate the effectiveness of AI models in classifying optimal versus suboptimal CXRs. Data was sourced from a large tertiary health network covering a range of specialties, including a dedicated children’s hospital. A substantial dataset of CXRs was collected, prepared, and divided for training, validation, and evaluation, with both models and radiologists participating in the assessment.

### Data preparation

Initially, 10,000 CXRs were retrieved from Picture Archiving and Communication System (PACS) servers using a Python script and the DICOM protocol. A randomised subset was then selected for training and validation. For model training, 4,000 suboptimal images were obtained from the Reject PACS, and 4,000 optimal images were obtained from the Diagnostic Imaging PACS. For validation, 400 optimal images and 400 suboptimal images were used. For evaluation, a radiographer with over 10 years of experience carefully selected 25 optimal and 25 suboptimal images from the remaining dataset, ensuring sufficient variance in patient demographics, rejection reasons, and image quality, including borderline cases. These images were then assessed by both the models and radiologists (Figures 1 and 2). Prior studies showed that test sets for different feature categories were roughly equivalent despite notable discrepancies in the volume of training data^9^. Consequently, this study applied a comparable method, using equivalent amounts of optimal and suboptimal test images.

**Figure 1.**
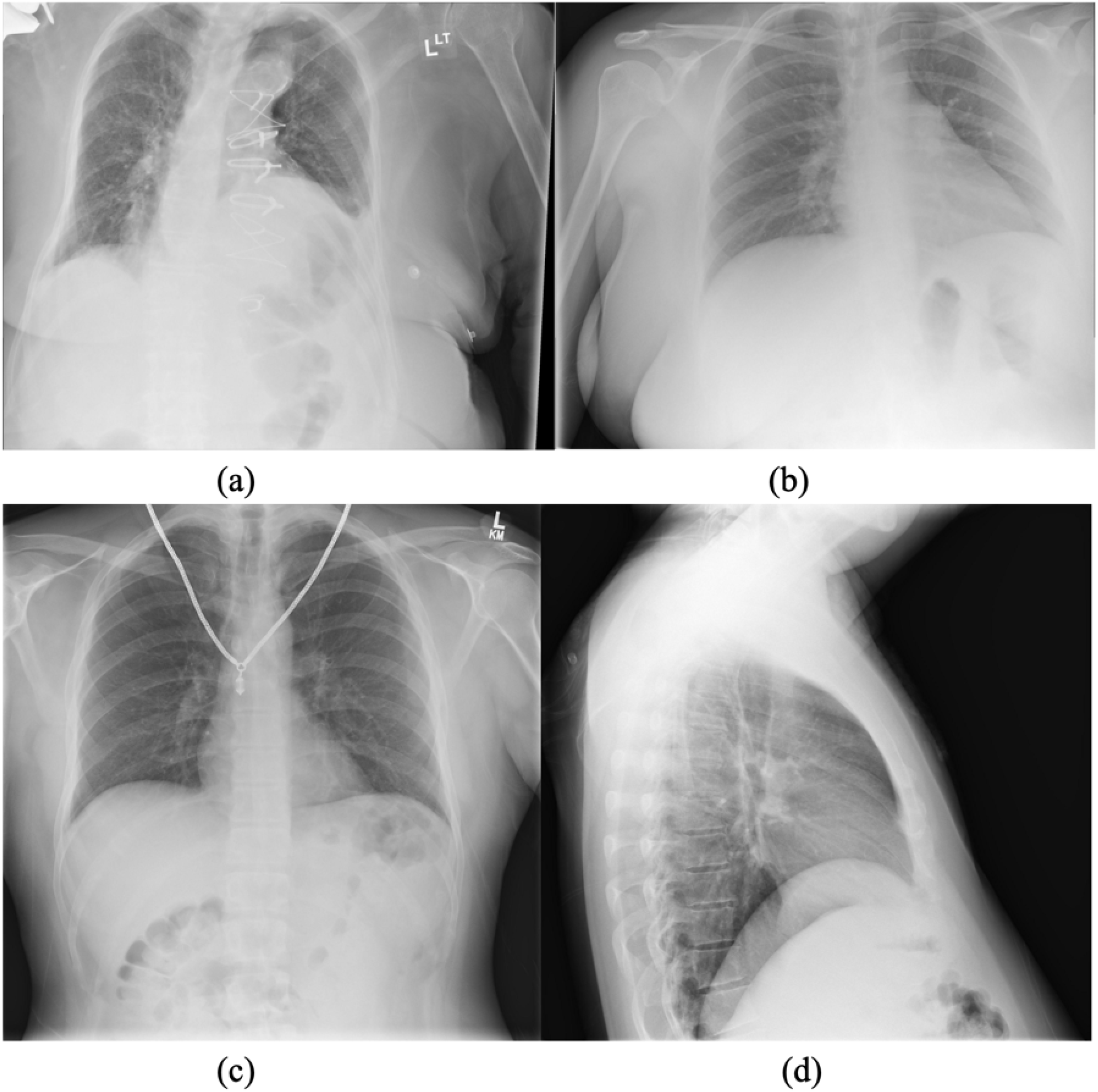
Rejected or suboptimal images with technical defects listed. (a) Right lung apices excluded (b) Patient motion and apices excluded (c) Necklace as artefact (d) Patient rotated and base of the lungs are excluded.

**Figure 2.**
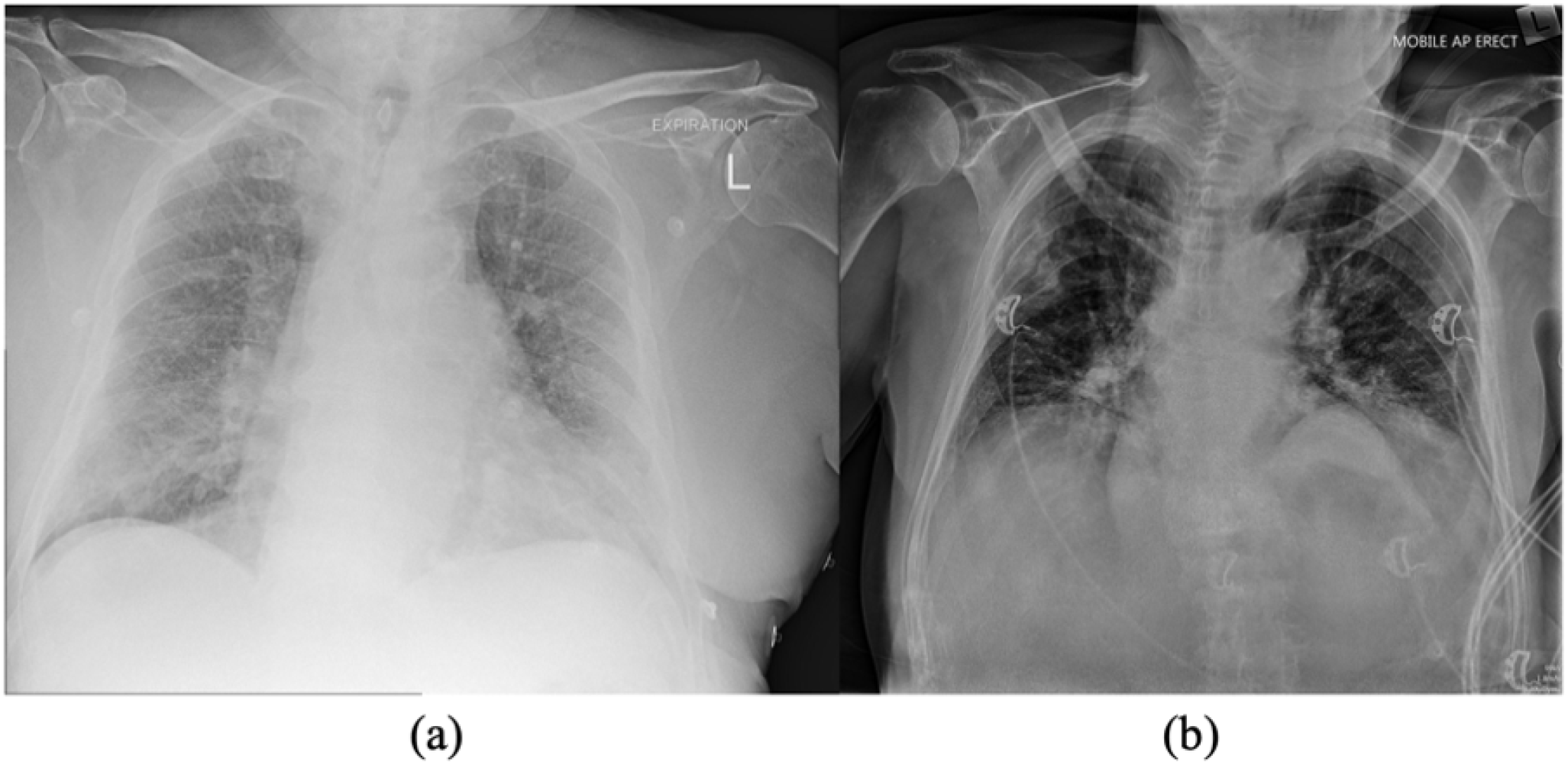
Accepted or optimal images and reasons for not repeating. (a) Poor exposure is accepted as all anatomy is included. (b) Inpatient CXR is accepted as lung markings are visualised.

The data retrieval method utilised convenience sampling without accounting for variables such as patient age, race, sex, radiographic equipment, or projection type. The images were limited to posteroanterior (PA), anteroposterior (AP), and lateral projections. They were resized to a resolution of 256×256 for optimal training on constrained computing resources. All study identifiers and corresponding images underwent anonymisation during the data extraction process, with no metadata preserved. The anonymised images were securely stored within a secure research computing environment for subsequent processing.

### Model training

The models underwent training for 200 epochs using PyTorch version 2.4 with Python 3.10. Training was performed on an NVIDIA P40 GPU with 24 GB of VRAM, using CUDA version 12.1. Hyperparameters such as batch size and learning rate were systematically adjusted to optimise training and validation outcomes. Weights and Biases (WandB) was used to visualise the training progression, and loss graphs were generated to assess model stability.

### Data analysis

The study utilised AUROC curves to assess the accuracy of AI models in image quality classification^8,9,11^. It was assumed that the neural network exhibited better performance as the AUROC approached 1.0^13^. When the AUROC exceeded 0.5, the test demonstrated significance, and a value of 0.8 or higher indicated that the classification was acceptable^14^.

Three radiologists, each possessing 5-10 years of expertise in the field, conducted a blind classification of the supplied images, designating them as either “No major technical deficiency” (optimal) or “Technical deficiency present” (suboptimal). The images were delivered as 256×256 JPEG files and presented to the radiologists through a Google Forms survey (Figure 3).

**Figure 3.**
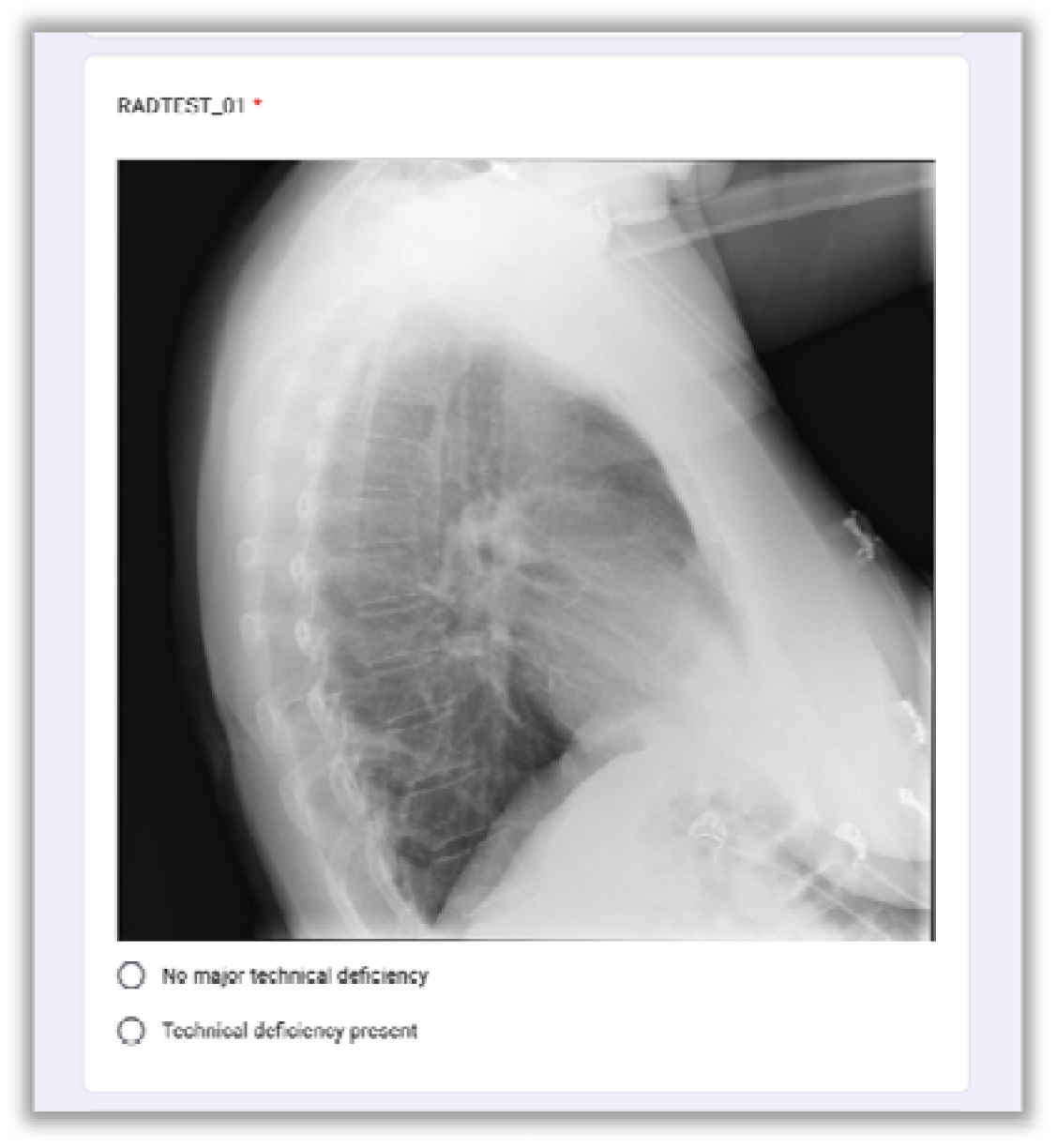
Screenshot of Google Form survey question provided to radiologists.

The classifications were gathered and analysed to evaluate the results of the two models and radiologists. A chi-squared test was used to determine whether the observed differences between the model classifications and those of the radiologists were statistically significant, indicating a genuine relationship rather than chance^15^. The goal was to assess whether the models’ classifications were comparable to those of the radiologists. Statistical analysis was conducted using the Python scikit-learn package.

### Ethics

The Ethics Review Board provided an exemption, classifying the study as a quality improvement project. This project was performed retrospectively with anonymised data, so informed consent was not required.

## Results

The patient cohort comprised 3587 individuals of adults and paediatrics, with a median age of 67 years, spanning a range from 0 to 102 years (Figure 4). The sample comprised 1986 males and 1601 females, providing a varied representation for model training and testing.

**Figure 4.**
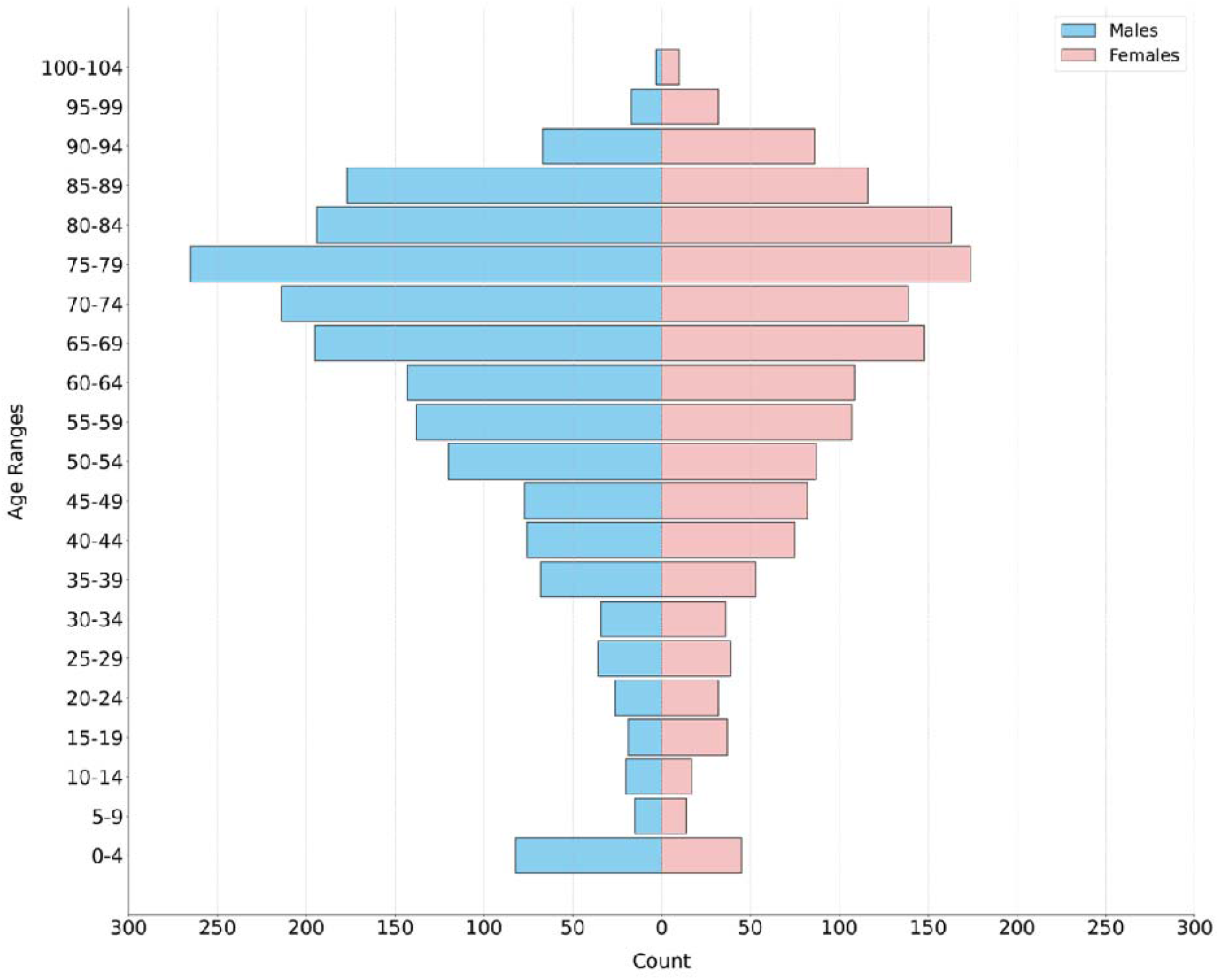
Frequency distribution by age and sex

Figures 5 and 6 showed a positive trend in the accuracy of YOLOv8 and DenseNet121, along with a decrease in training and validation loss that eventually stabilises^16^. The YOLOv8 loss curves suggest potential for further improvement if left to run for longer; however, they were kept at 200 epochs for consistency.

**Figure 5.**
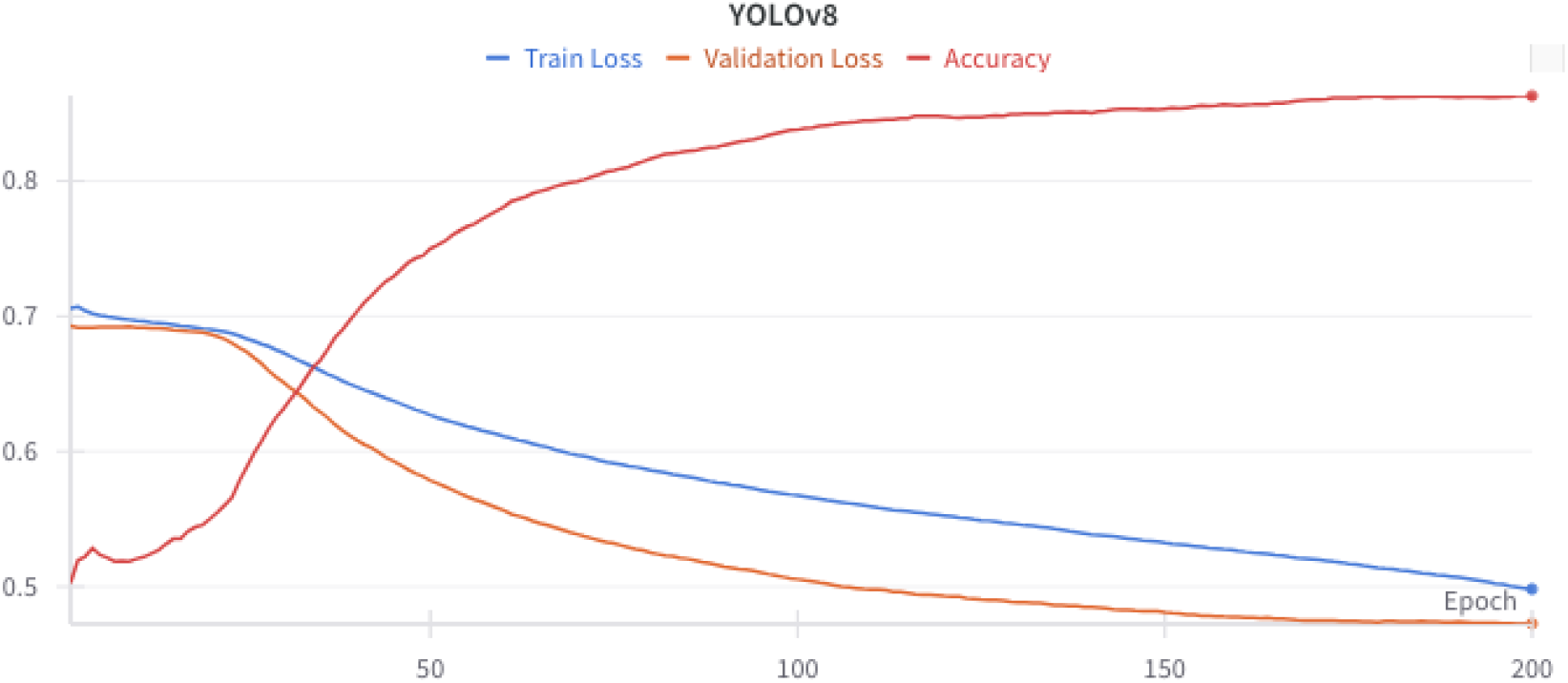
Training loss (blue), validation loss (orange) and accuracy (red) of YOLOv8 network during training.

**Figure 6.**
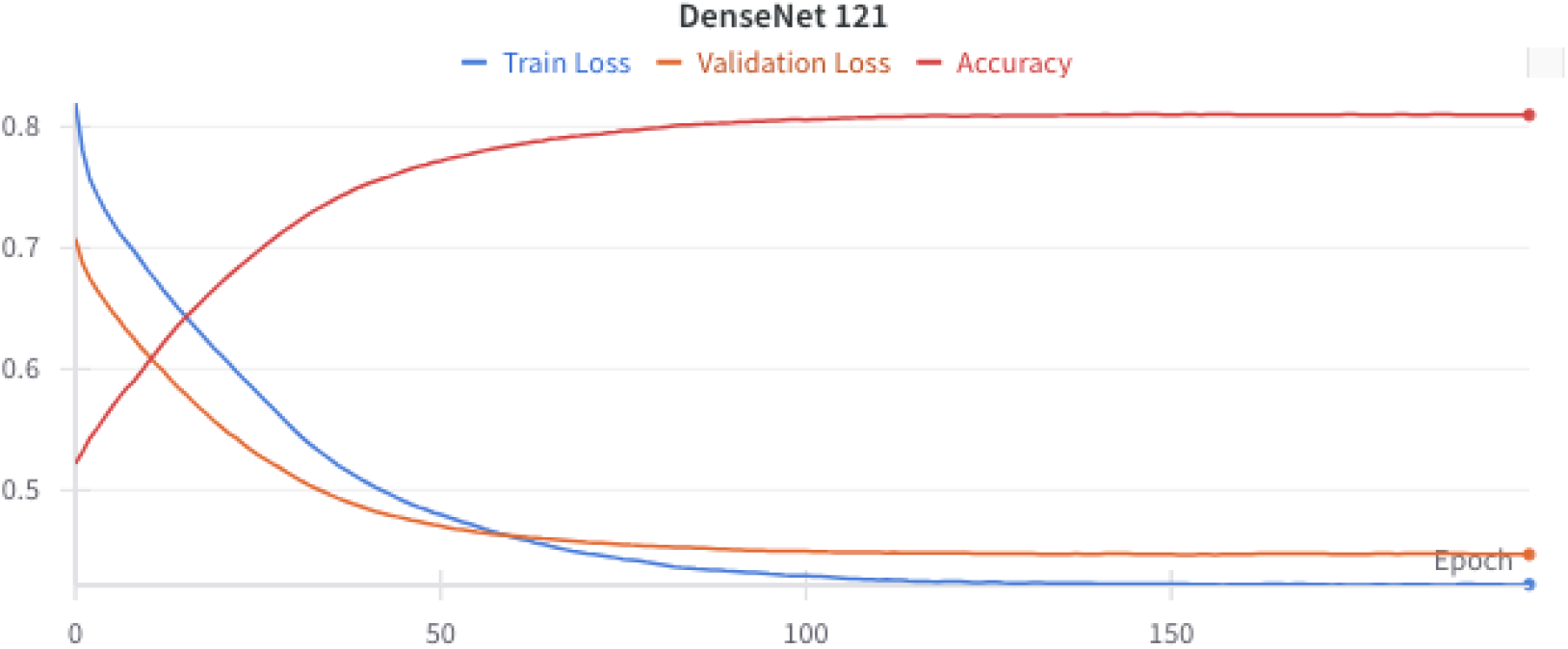
Training loss (blue) vs validation loss (orange) and accuracy (red) of DenseNet121 network during training.

The DenseNet121 network correctly predicted 24 (96%) of optimal and 20 (80%) of suboptimal images (Figure 7). The ROC curve of the model’s performance was plotted with an AUC of 0.97. YOLOv8 correctly predicted 92% of optimal images and 88% of suboptimal images with an AUC of 0.95 (Figure 8).

**Figure 7.**
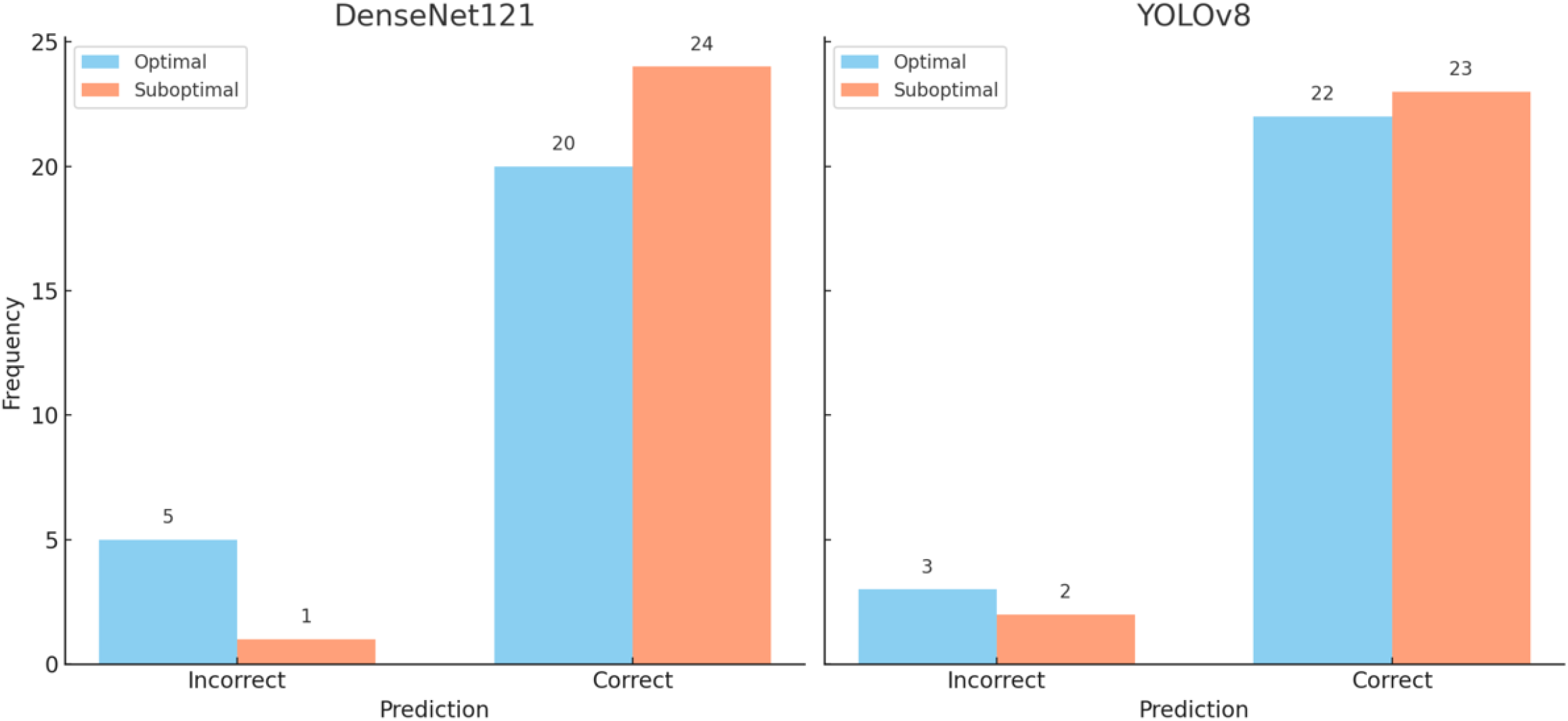
Prediction of optimal vs suboptimal CXRs by DenseNet121 and YOLOv8

**Figure 8.**
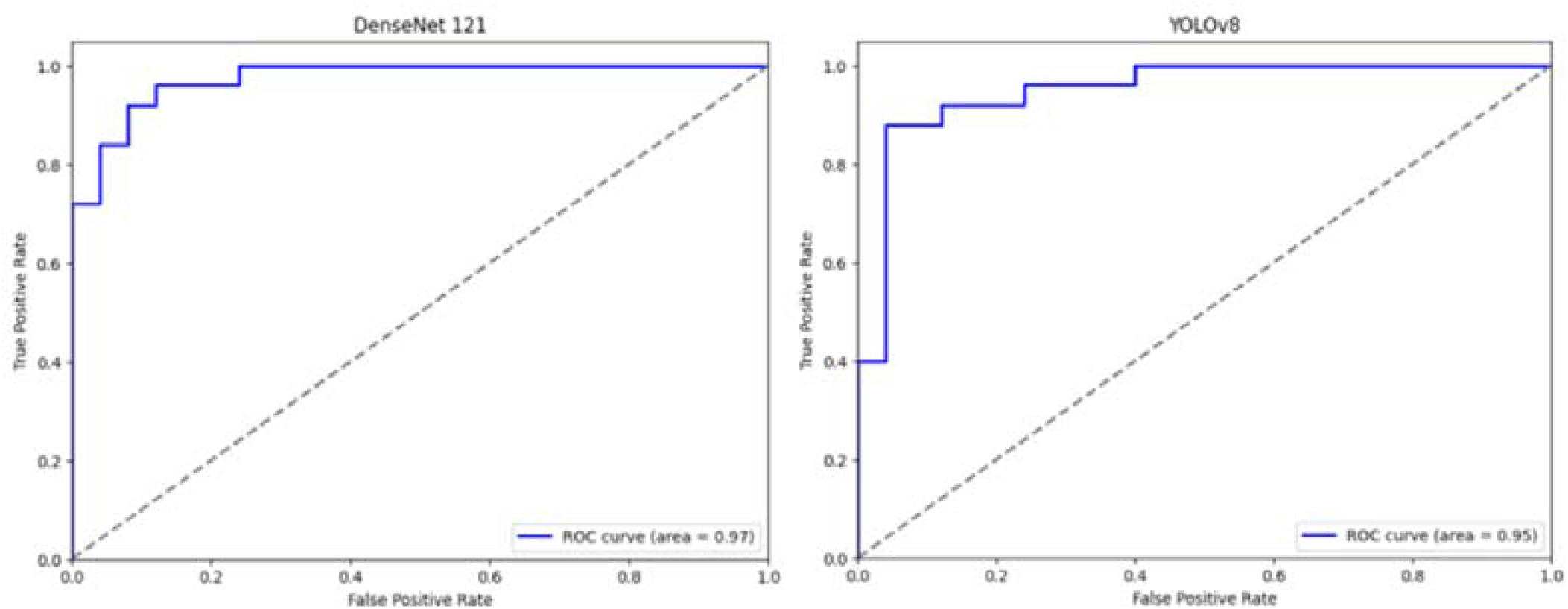
ROC curve of DenseNet121 vs YOLOv8’s performance

A precision-recall graph was created to compare the models and radiologists in classifying optimal and suboptimal images. Precision refers to the classifications corresponding with the categories of optimal and suboptimal images. Recall indicates the proportion of optimal images successfully identified by the models. DenseNet121 showed the highest precision at 0.95, indicating accurate identification of both categories, while its recall was slightly lower at 0.8, reflecting some missed optimal images. YOLOv8 demonstrated a balanced precision of 0.9 and a higher recall of 0.85, suggesting better identification of optimal images with slightly reduced precision. The radiologists had varying precision and recall scores, indicating differing levels of accuracy and consistency, with Radiologist 3 having the highest recall among them (Figure 9).

**Figure 9.**
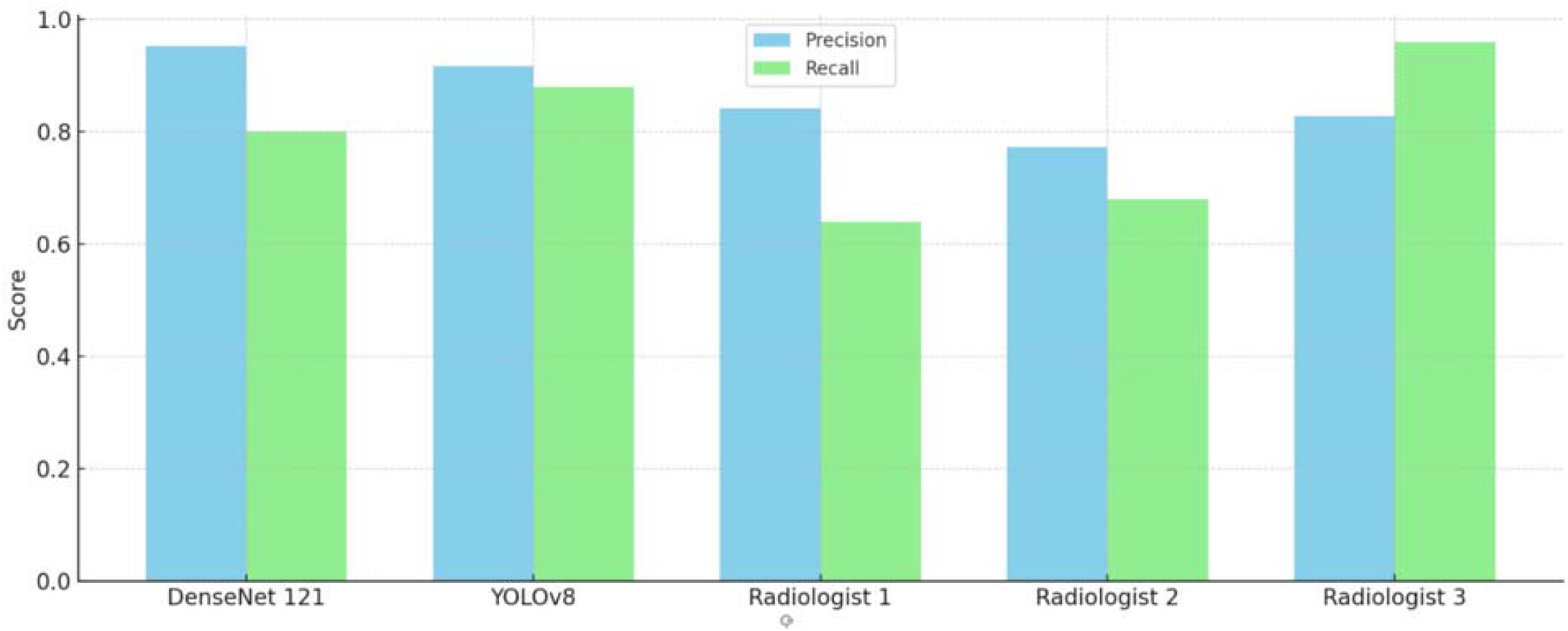
Precision and recall of models and radiologists.

The differences between the radiologists and the models exhibit some variance, however, these differences are non-statistically significant, suggesting comparable performance across all methods. Although there is some variability in the radiologists’ performance, the p-values for both models show no significant difference when compared to the combined confusion matrices of radiologists’, implying similar levels of effectiveness (Figure 10).

**Figure 10.**
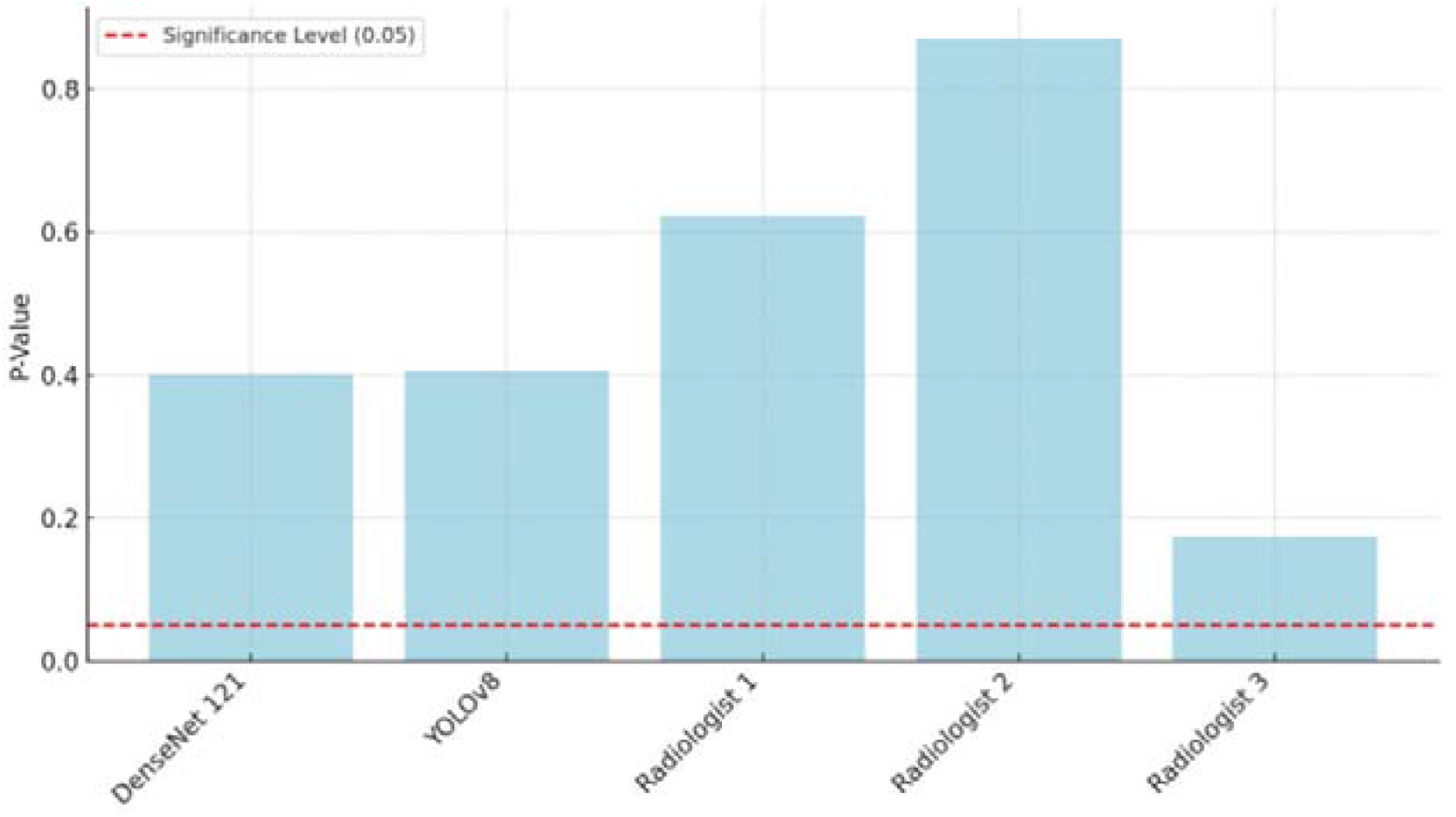
Chi-squared test of DenseNet121, YOLOv8 and individual radiologists vs the combined confusion matrices of radiologists.

**Figure 11.**
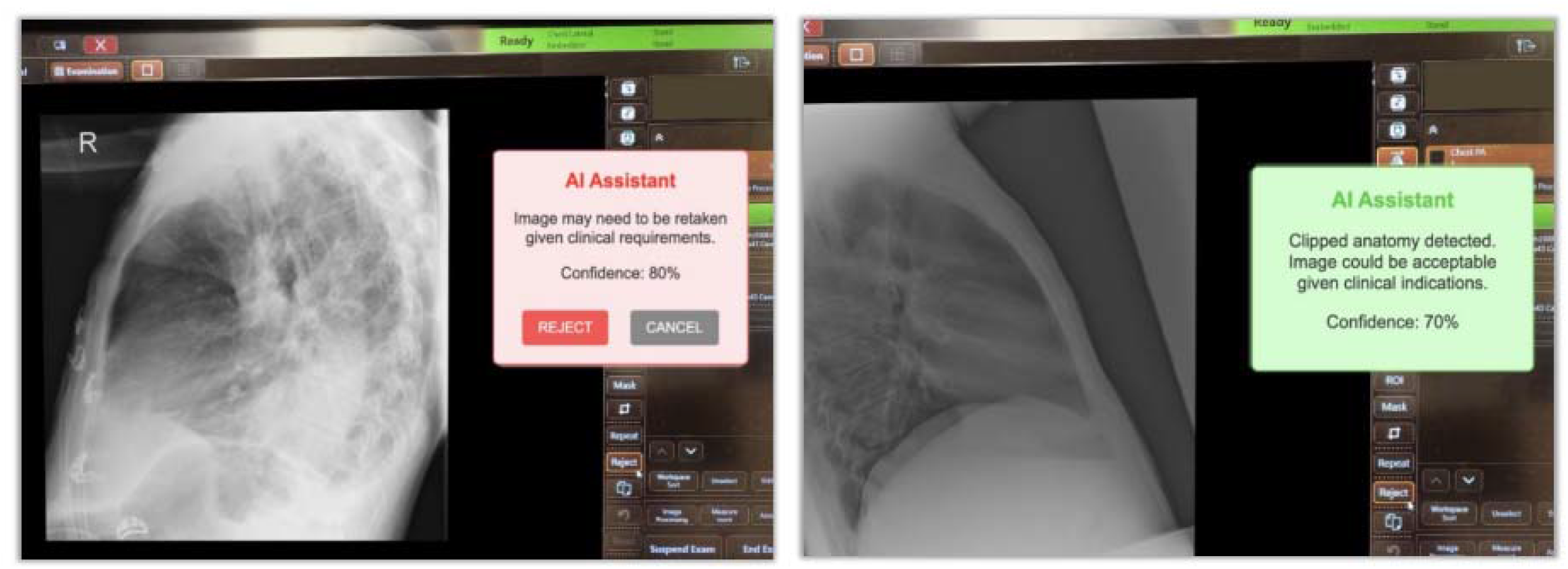
Example proof of concept AI Assisted Feedback at an X-ray console. The left image indicates that the X-ray requires repeating, while the right image suggests that it may still be acceptable considering the clinical context.

## Discussion

This study’s results indicated that CNNs could be trained to detect suboptimal CXRs. Both models’ ROC curves showed outstanding performance, with AUC measuring 0.97 for DenseNet121 and 0.95 for YOLOv8, suggesting strong classification ability while minimising false positives. DenseNet121 slightly outperformed YOLOv8, yet both models demonstrated potential for clinical application with opportunities for further refinement. Notably, this was the first study to include both PA and lateral chest X-rays, as well as pediatric cases, and to test YOLOv8 in this context. The consistent performance of the CNNs across diverse patient age groups, sexes, and radiographic projections highlights their robustness in image quality classification.

Research in related domains had similarly demonstrated the efficacy of DenseNet121 in identifying suboptimal X-rays with an AUC of 0.93^6^. In a separate study, the performance of AI models was divided into subcategories based on technical deficiencies in images. The models obtained 0.87 AUC for missing anatomy, 0.91 for inadequate exposure, 0.94 for low lung volume, and 0.94 for patient rotation^7^. A direct comparison with the second study was not feasible as our research did not include subclasses, which was not possible at the time of research due to resource constraints. While our model demonstrated superior performance on suboptimal images compared to findings from Kashyap et al.^8^, this discrepancy might be attributed to differences in the training and test datasets. Literature has shown the promising performance of YOLOv8 in detecting tumours, classifying diseases, and organ segmentation^11^. However, no other studies have investigated YOLOv8’s accuracy in identifying suboptimal-quality radiographs.

Early identification of suboptimal images offers significant benefits, such as reducing the need for repeat imaging, which in turn minimises radiation exposure to patients. Additionally, fewer imaging attempts reduce repositioning time and alleviate patient discomfort. However, integrating deep learning models into healthcare systems presents several challenges, including issues with reproducibility, limited explainability of model outputs, potential unintended biases, privacy concerns, compliance requirements, and increased costs. Therefore, thorough validation is crucial to prevent unexpected model failures^13^. A more practical implementation could involve auditing all CXRs sent to PACS, where the models would flag suboptimal images and provide a feedback loop for training and quality improvement.

### Future work and applications

Future research may benefit from integrating clinical details before model training, which could have enriched the clinical context and enhanced both model accuracy and radiologists’ image quality assessment. Suboptimal X-rays could also be classified into subclasses during training, enabling the model to recognise rejection reasons, such as anatomical exclusion, improper exposure, poor inspiration, mispositioning, and overlying anatomy^6^. Additionally, involving radiologists in grading the dataset before training could yield a more dependable ground truth.

Figure 9 presents a mocked implementation of a flagging tool designed to enhance the application of AI in radiographic practice. Such flagging tools could significantly improve educational practices and facilitate quality audits. Integrating a model capable of flagging technical deficiencies directly at the imaging console offers the potential for immediate feedback to radiographers following X-ray acquisition. Intervention could be particularly beneficial for students, medical imaging interns, and junior radiographers, who often face challenges accepting or rejecting suboptimal X-rays. In this context, AI models could provide real-time feedback, enhancing junior staff’s decision-making skills. Importantly, the patient’s clinical indications should inform the decision to accept or repeat an X-ray.

Lastly, these models can be an effective auditing tool for large image datasets. Through retrospective audits, they may help identify X-rays that require quality improvements, thereby uncovering opportunities for additional training or enhancing adherence to protocols.

This approach eliminates the need to manually curate rejected data, which is time-consuming and laborious.

### Limitations

This research presents some limitations. Firstly, images were categorised into optimal and suboptimal based on where they were delivered. All images transmitted to Diagnostic Imaging PACS were considered optimal, whereas those directed to Reject PACS were classified as suboptimal. The classification relied exclusively on the radiographers’ judgement, without any prior classification by radiologists, who are regarded as the benchmark for evaluating image quality. The dependence on the judgement of radiographers may have influenced the precision of the reference standard employed in this study.

Furthermore, the absence of clinical indications within the dataset led to inconsistencies in the radiologist image quality assessment. Without comprehensive clinical information, the models and radiologists encountered difficulties in consistently evaluating the quality of the CXRs. This limitation reduced inter-observer agreement among radiologists, thereby impacting the overall reliability of the assessments.

Lastly, the dataset encompassed inpatient and outpatient cases, introducing variability in the necessary image quality standards. Inpatient imaging frequently accommodates lower quality standards due to patients’ critical conditions and clinical requirements, while outpatient imaging necessitates higher quality to facilitate precise diagnoses. The differences in image quality standards between inpatients and outpatients presented a challenge for model training, complicating the model’s ability to generalise across various clinical settings. It is essential to consider these limitations when analysing the study’s findings and assessing the broader relevance of the model.

## Conclusion

This research demonstrated that DenseNet121 and YOLOv8 effectively categorise CXRs into optimal and suboptimal classes, achieving AUROC scores of 0.97 and 0.95, respectively. These models show significant potential for auditing radiographic quality, which may reduce the need for repeat imaging, lower patient radiation exposure, and improve workflow efficiency in clinical practice.

Future research should focus on refining these models to identify specific technical errors and incorporate clinical details for increased contextual precision. AI-assisted feedback tools show promise for providing real-time support to radiographers, while automated quality control systems could standardise imaging practices and improve patient outcomes. These findings highlight the potential of AI to enhance radiographic quality and diagnostic accuracy.

## Data Availability

All data produced in the present study are available upon reasonable request to the authors.

## Notes

### Competing Interest Statement

The authors have declared no competing interest.

### Funding Statement

This study did not receive any funding.

### Author Declarations

Ethics committee of Monash Health gave ethical approval for this work.

## References

1. Shakya DrS. Analysis of Artificial Intelligence based Image Classification Techniques. JIIP. 2020;2(1):44–54. doi:10.36548/jiip.2020.1.005

2. Hosny A, Parmar C, Quackenbush J, Schwartz LH, Aerts HJWL. Artificial intelligence in radiology. Nat Rev Cancer. 2018;18(8):500–510. doi:10.1038/s41568-018-0016-5

3. Meng Y, Ruan J, Yang B, et al. Automated quality assessment of chest radiographs based on deep learning and linear regression cascade algorithms. Eur Radiol. 2022;32(11):7680–7690. doi:10.1007/s00330-022-08771-x

4. Calatayud-Jordán J, Campayo-Esteban JM, Gras-Miralles P, Villaescusa-Blanca JI. Image reject analysis and rejection causes in digital radiography at a university hospital through estimates of patient doses. Radiation Protection Dosimetry. 2024;200(3):274–284. doi:10.1093/rpd/ncad301

5. Mount J. Reject analysis: A comparison of radiographer and radiologist perceptions of image quality. Radiography. 2016;22(2):e112–e117. doi:10.1016/j.radi.2015.12.001

6. Dasegowda G, Bizzo BC, Gupta RV, et al. Radiologist-Trained AI Model for Identifying Suboptimal Chest-Radiographs. Academic Radiology. 2023;30(12):2921–2930. doi:10.1016/j.acra.2023.03.006

7. Kjelle E, Schanche AK, Hafskjold L. To keep or reject, that is the question - A survey on radiologists and radiographers’ assessments of plain radiography images. Radiography. 2021;27(1):115–119. doi:10.1016/j.radi.2020.06.020

8. Moradi M, Siegel E, Kashyap S, et al. Artificial intelligence for point of care radiograph quality assessment. In: Hahn HK, Mori K, eds. Medical Imaging 2019: Computer-Aided Diagnosis. SPIE; 2019:128. doi:10.1117/12.2513092

9. Nousiainen K, Mäkelä T, Piilonen A, Peltonen JI. Automating chest radiograph imaging quality control. Physica Medica. 2021;83:138–145. doi:10.1016/j.ejmp.2021.03.014

10. Huang G, Liu Z, Maaten L van der, Weinberger KQ. Densely Connected Convolutional Networks. Published online January 28, 2018. Accessed November 10, 2024. http://arxiv.org/abs/1608.06993

11. Widayani A, Putra AM, Maghriebi AR, Adi DZC, Ridho MohHF. Review of Application YOLOv8 in Medical Imaging. IAPL. 2024;5(1):23–33. doi:10.20473/iapl.v5i1.57001

12. Dasegowda G, Kalra MK, Abi-Ghanem AS, et al. Suboptimal Chest Radiography and Artificial Intelligence: The Problem and the Solution. Diagnostics. 2023;13(3):412. doi:10.3390/diagnostics13030412

13. Yang HS, Rhoads DD, Sepulveda J, Zang C, Chadburn A, Wang F. Building the Model. Archives of Pathology & Laboratory Medicine. 2023;147(7):826–836. doi:10.5858/arpa.2021-0635-RA

14. Nahm FS. Receiver operating characteristic curve: overview and practical use for clinicians. Korean J Anesthesiol. 2022;75(1):25–36. doi:10.4097/kja.21209

15. Kim HY. Statistical notes for clinical researchers: Chi-squared test and Fisher’s exact test. Restor Dent Endod. 2017;42(2):152. doi:10.5395/rde.2017.42.2.152

16. Smith LN. A disciplined approach to neural network hyper-parameters: Part 1 -- learning rate, batch size, momentum, and weight decay. Published online 2018. doi:10.48550/ARXIV.1803.09820

